# A case-crossover study of the effect of vaccination on SARS-CoV-2 transmission relevant behaviours during a period of national lockdown in England and Wales

**DOI:** 10.1101/2022.08.29.22279333

**Authors:** Aimee Serisier, Sarah Beale, Yamina Boukari, Susan Hoskins, Vincent Nguyen, Thomas Byrne, Wing Lam Erica Fong, Ellen Fragaszy, Cyril Geismar, Jana Kovar, Alexei Yavlinsky, Andrew Hayward, Robert W Aldridge

## Abstract

**Background:** Studies of COVID-19 vaccine effectiveness show increases in COVID-19 cases within 14 days of a first dose, potentially reflecting post-vaccination behaviour changes associated with SARS-CoV-2 transmission before vaccine protection. However, direct evidence for a relationship between vaccination and behaviour is lacking. We aimed to examine the association between vaccination status and self-reported non-household contacts and non-essential activities during a national lockdown in England and Wales.

**Methods:** Participants (n=1,154) who had received the first dose of a COVID-19 vaccine reported non-household contacts and non-essential activities from February to March 2021 in monthly surveys during a national lockdown in England and Wales. We used a case-crossover study design and conditional logistic regression to examine the association between vaccination status (pre-vaccination vs. 14 days post-vaccination) and self-reported contacts and activities within individuals. Stratified subgroup analyses examined potential effect heterogeneity by sociodemographic characteristics such as sex, household income or age group.

**Results:** 457/1,154 (39.60%) participants reported non-household contacts post-vaccination compared with 371/1,154 (32.15%) participants pre-vaccination. 100/1,154 (8.67%) participants reported use of non-essential shops or services post-vaccination compared with 74/1,154 (6.41%) participants pre-vaccination. Post-vaccination status was associated with increased odds of reporting non-household contacts (OR 1.65, 95% CI 1.31-2.06, p<0.001) and use of non-essential shops or services (OR 1.50, 95% CI 1.03-2.17, p=0.032). This effect varied between men and women and different age groups.

**Conclusion:** Participants had higher odds of reporting non-household contacts and use of non-essential shops or services within 14 days of their first COVID-19 vaccine compared to pre-vaccination. Public health emphasis on maintaining protective behaviours during this post-vaccination time period when individuals have yet to develop full protection from vaccination could reduce risk of SARS-CoV-2 infection.

## INTRODUCTION

The coronavirus disease 2019 (COVID-19) pandemic has had a devastating impact on global public health since the causative agent, severe acute respiratory syndrome coronavirus 2 (SARS-CoV-2) was first identified in late 2019 (1,2). SARS-CoV-2 is transmitted through direct or indirect contact with infected respiratory droplets or aerosols (3–5). Consequently, public settings and activities that involve direct and indirect contact may promote SARS-CoV-2 transmission (4,6) and were the target of non-pharmaceutical interventions (NPIs) introduced by many governments worldwide to control the spread of the virus. COVID-19 vaccination programmes, which began in December 2020 with the emergency licencing of first the Pfizer BioNTech and then AstraZeneca vaccines in the UK, are now a cornerstone of pandemic response in the UK and worldwide along with varying degrees of remaining NPIs. Delivered as two doses with additional booster doses, first and subsequent doses are proven to be effective in reducing symptomatic and asymptomatic infections, hospitalisations, and deaths from COVID-19 (7–10). However, the protection provided by vaccines is not immediate and infection with SARS-CoV-2 after vaccination is possible (11). Studies using data from Israel and the UK suggest an increased risk of symptomatic infection within 14 days of the first dose of a COVID-19 vaccine (12,13); the reduction in risk of symptomatic infection is seen after 14 days. It is possible that increased risk in the 14 days after vaccination may reflect changes in behaviour associated with SARS-CoV-2 transmission during the period of time in which immunologic protection is building (14-16), particularly given the extent to which human behaviour is known to influence infectious disease dynamics (17-20). Furthermore, concerns have been raised that a reduction in behaviours protecting against SARS-CoV-2 infection could be seen if the perceived risk of infection is reduced after vaccination against COVID-19 (21).

Empirical evidence regarding the effect of vaccination on infection prevention behaviour is limited. One study examining protective behaviours before and after vaccination against Lyme disease found that people who were vaccinated reduced protective behaviours and believed that they were at less risk of infection than unvaccinated people (22). There is evidence in the context of the COVID-19 pandemic that suggests changes in protective behaviours may occur. In February 2021 the Office for National Statistics (ONS) reported that 41% of over 80s met a person who was not a part of their household or support bubble indoors within 21 days post-vaccination (23). A December 2020 YouGov survey poll found that of the 1,706 people surveyed, 29% said that they would follow public health restrictions less strictly after receiving a vaccine (24); this poll was conducted prior to widespread availability of COVID-19 vaccination in the UK. Reductions in compliance with mask use and handwashing post-vaccination were found amongst healthcare workers in Ethiopia after the first dose of a COVID-19 vaccine (25). Contradictory to these findings, a longitudinal analysis investigating compliance with protective behaviours found increases in self-reported compliance with public health guidance and social distancing in vaccinated and unvaccinated individuals from October 2020 to February 2021 (26). However, variation in context over this time could have influenced these results - for example, the introduction of the third national lockdown in January 2021 or changes in the perceived risk of infection as case numbers rose.

This study aimed to quantify the effect of a change in COVID-19 vaccination status on transmission-relevant behaviours during a period of national lockdown using data from Virus Watch, a large prospective cohort study based in England and Wales. We set out to investigate whether participants’ self-reported levels of non-household contacts and retail and social activities classed as non-essential under lockdown restrictions (such as use of a hairdressers or other services for personal care, attending a party, or dining at restaurant or café) changed within 14 days of their first dose of a COVID-19 vaccination compared to pre-vaccination.

## METHODS

### Study design

A case-crossover design was used to examine the association between vaccination status (pre-vaccination versus ≤14 days post first dose) and self-reported contacts and activities within individuals. Case-crossover designs are appropriate to examine the association between transient exposures and acute outcomes (27), and eliminate measured and unmeasured time-invariant confounding when comparing within-person exposed and unexposed periods (27-30).

To maximise the comparability of referent periods and reduce the potential for confounding by temporal or spatial trends in contacts and activities, the study timeframe was limited to include survey responses from within the third national lockdown in England and Wales (6 January – 29 March 2021). Each survey was open for 7 days and concerned non-household contacts and activities in the week prior to its start date – surveys from 9-16 February 2021 and 9-16 March 2021 could therefore be included, and the pre-vaccination period comprised responses from 2-9 February 2021 and the post-vaccination period from 2-9 March 2021.

### Study setting and population

Data analysed in this project were collected as part of Virus Watch, a household community cohort study of SARS-CoV-2 transmission and COVID-19 in England and Wales. Details of Virus Watch relevant to the present study are described briefly here, with further information on its full scope and methodology described in the study protocol (31).

Participants were recruited in households, and a baseline survey completed upon study registration collected information on sociodemographic factors for each household member. Participants reported on changes in the vaccination status weekly. Monthly surveys collected health-related and behavioural/psychosocial factors, including information about participants’ activities and contacts.

Inclusion criteria were having received the first dose of a COVID-19 vaccine and responded to a monthly survey regarding activities and contacts both pre-vaccination and post-vaccination (n = 1,154). Participants under 18 years old were excluded (n = 4) as this age group was not the focus of vaccinations during the study timeframe. Participants with missing data on sociodemographic covariates (see below) to be used in stratified analyses were excluded from the study sample following initial descriptive statistics (n = 23). The final study population for analysis included 1,141 participants (Figure 1).

### Exposure

Vaccination status was the exposure of interest, defined as ‘pre-vaccination’ and ‘within 14-days post-vaccination’ of a first dose of a COVID-19 vaccine and derived from data on the date and dose of vaccinations reported in weekly surveys.

### Outcome

The following contact and activity outcomes were derived based on responses to monthly surveys, and were binary coded (yes or no during survey week): (1) any close contact with non-household members (‘face-to-face contact with another person less than 1 metre away, spending more than 15 minutes within 2 metres of another person, or travelling in a car or other small vehicle with another person’ (32), (2) any social or leisure activity (attending a theatre, cinema, concert, or sports event; attending a party; going to a restaurant, café, or canteen; going to a bar, pub, or club; and/or use of a gym or indoor sports facility), and (3) visiting non-essential shops or services (e.g. hairdressers, barbers, or beauty salons). Such behaviours were targeted by social distancing restrictions and/or public venue closures under lockdown restrictions in place in February – March 2021. Due to low numbers of participants reporting social activities in both pre- and post-vaccination surveys during this lockdown period, social and leisure activities could not be included in this analysis.

### Demographic Characteristics

Data on sociodemographic characteristics were collected from study baseline responses. Age group was defined according to reported age of participants at baseline: less than 60 years and 60 years or more. Participants were categorised as White British and Minority Ethnic according to self-reported ethnicity. Sex was categorised as male and female by self-reported sex. Household size was defined as households of 1 person, 2-3 people and 4-6 people. The Index of Multiple Deprivation (IMD) for each participant was categorised using household postcodes recorded on registration, comprising 1 (least deprived) through 5 (most deprived). Region was measured using participant’s postcodes and categorised according to ONS national regions. Observations with missing data on sex (1.12%) and age (0.43%) which were included as potential effect modifiers were excluded following descriptive statistics in order to perform a complete cases analysis.

### Analysis

The frequency with which non-household contacts and use of non-essential shops or services were reported in surveys pre- and post-vaccination were calculated and stratified by age and sex only as group sizes amongst other participant subgroups were too small. Conditional logistic regression was used to assess the odds of reporting contacts and activity outcomes within 14 days after the first dose of a COVID-19 vaccine compared to the earlier pre-vaccination time point within individuals. This approach accounts for non-independence in responses within individuals over time by conditioning the participants responses using an individual-specific fixed effect (33). Each participant contributes two observations occupying a single stratum in the model. As the model estimates within-individual changes, strata that do not vary (i.e., people who reported contacts and activities in neither hazard nor control periods, or in both) do not contribute to the model (29). Stratification of conditional logistic regression models by age and sex was then performed. Sociodemographic covariates were collected at baseline and considered time-invariant confounders, and consequently were not entered into the models. Cluster robust standard errors were used to account for household-level clustering.

### Ethics

Ethical approval for Virus Watch was obtained from the Hampstead National Health Service (NHS) Health Research Authority Ethics Committee (ethics approval number 20/HRA/2320). All participants provided online informed consent. All analyses were conducted using the University College London Data Safe Haven.

## RESULTS

Table 1 presents the sociodemographic characteristics of Virus Watch participants in this analysis (n = 1,154). The majority were over 60 years of age (58.12%), likely due to the phases of COVID-19 vaccine delivery, with older age groups being prioritised in the early stages of the COVID-19 vaccination programme (34). Participants were mostly white (95.93%), from the East of England (24.61%) or South East (20.10%), lived in households with 2-3 people (62.48%), in IMD 5 (least deprived, (31.63%) or with an annual household income of £50,000 or more (38.88%). There was a greater proportion of men in the sample (54.59%) compared to women (44.37%).

**Table 1.**
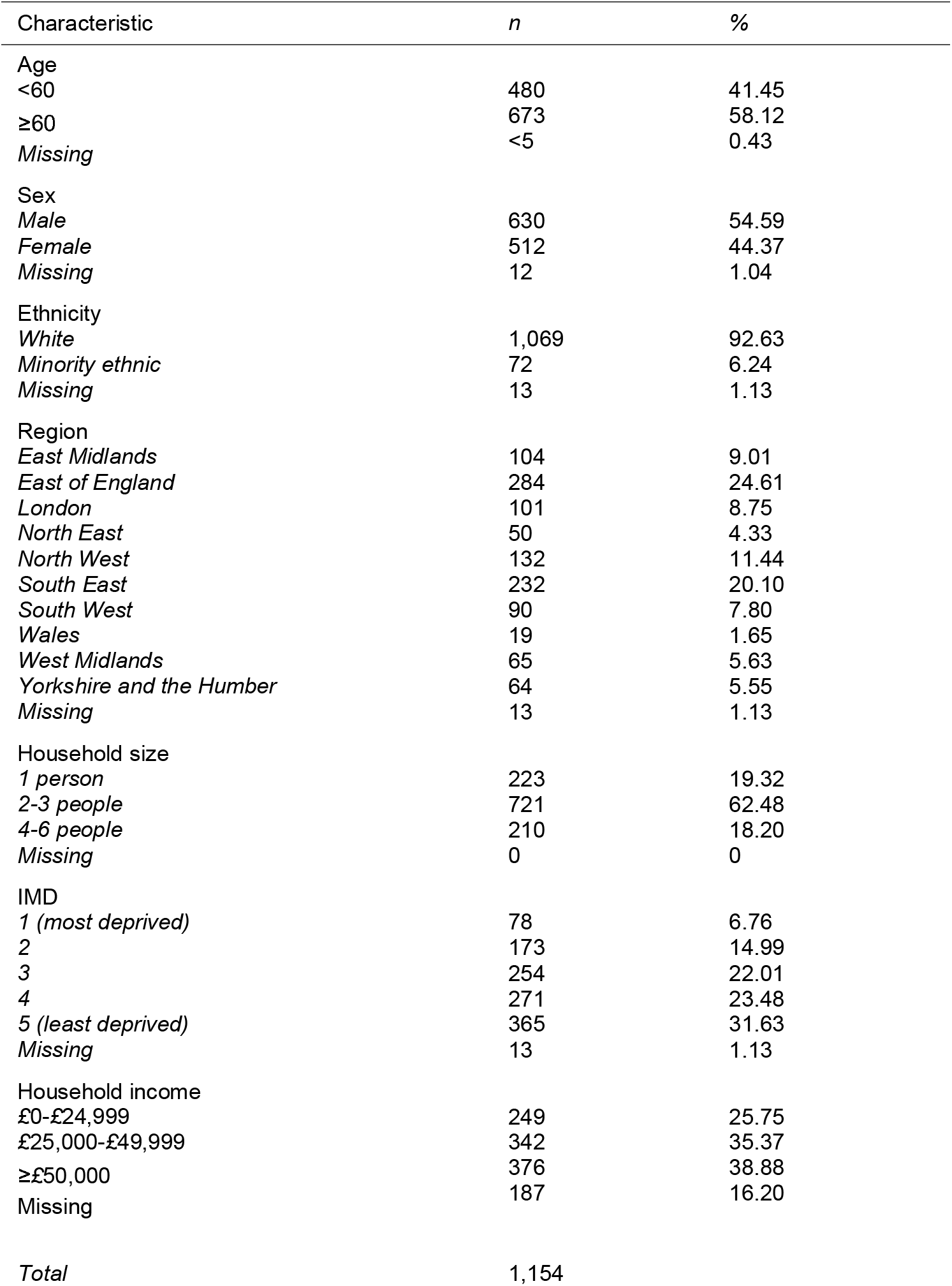
Characteristics of study participants

| Characteristic | *n* | % |
| --- | --- | --- |
| Age |  |  |
| <60 | 480 | 41.45 |
| ≥60 | 673 | 58.12 |
| Missing | <5 | 0.43 |
| Sex |  |  |
| Male | 630 | 54.59 |
| Female | 512 | 44.37 |
| Missing | 12 | 1.04 |
| Ethnicity |  |  |
| White | 1,069 | 92.63 |
| Minority ethnic | 72 | 6.24 |
| Missing | 13 | 1.13 |
| Region |  |  |
| East Midlands | 104 | 9.01 |
| East of England | 284 | 24.61 |
| London | 101 | 8.75 |
| North East | 50 | 4.33 |
| North West | 132 | 11.44 |
| South East | 232 | 20.10 |
| South West | 90 | 7.80 |
| Wales | 19 | 1.65 |
| West Midlands | 65 | 5.63 |
| Yorkshire and the Humber | 64 | 5.55 |
| Missing | 13 | 1.13 |
| Household size |  |  |
| 1 person | 223 | 19.32 |
| 2-3 people | 721 | 62.48 |
| 4-6 people | 210 | 18.20 |
| Missing | 0 | 0 |
| IMD |  |  |
| 1 (most deprived) | 78 | 6.76 |
| 2 | 173 | 14.99 |
| 3 | 254 | 22.01 |
| 4 | 271 | 23.48 |
| 5 (least deprived) | 365 | 31.63 |
| Missing | 13 | 1.13 |
| Household income |  |  |
| £0-£24,999 | 249 | 25.75 |
| £25,000-£49,999 | 342 | 35.37 |
| ≥£50,000 | 376 | 38.88 |
| Missing | 187 | 16.20 |
| Total | 1,154 |  |

Table 2 shows the frequency of reporting non-household contacts and use of non-essential shops or services. Pre-vaccination surveys asked participants to record activities from 2-9 February 2021, with post-vaccination surveys recording activities from 2-9 March 2021. 371/1,154 (32.15%) participants reported contact with a person outside of their household or support bubble pre-vaccination compared with 457/1,154 (39.60%) participants in the 14 days after the first dose of a COVID-19 vaccine. 74/1,154 (6.41%) participants reported using non-essential shops or services pre-vaccination, while 100/1,154 (8.67%) participants did so in the 14 days after their first COVID-19 vaccine.

**Table 2.** Frequency of reporting non-household contacts and use of non-essential shops or services over 7 days in pre- and post-vaccination surveys (n = 1,154).

| Activity | *Pre-vaccination* |  | *≤14 days post-vaccination* |  |
| --- | --- | --- | --- | --- |
|  | *No* | *Yes* | *No* | *Yes* |
|  | *n (%)* | *n (%)* | *n (%)* | *n (%)* |
| **Contacts** | 783 (67.85) | 371 (32.15) | 697 (60.40) | 457 (39.60) |
| **Use of non-essential shops or services** | 1,080 (93.59) | 74 (6.41) | 1,054 (91.33) | 100 (8.67) |

*The pre-vaccination survey and post-vaccination surveys asked participants to report activities from 2-9 February 2021 and 2-9 March 2021 respectively.*

Table 3 shows the frequency with which participants reported activities in pre-vaccination and post-vaccination surveys stratified by age group and sex. A greater proportion of participants reported non-household contacts in both pre-vaccination and post-vaccination surveys compared to use of non-essential shops and services, but both activities were reported more frequently post-vaccination. Post-vaccination, reporting of non-household contacts was greatest amongst male participants and those aged less than 60 (257/630, 40.79% and 199/480, 41.46% respectively), but the greatest increase in reported non-household contacts was seen when comparing post-vaccination with pre-vaccination surveys from male participants and those aged 60 years or more. Greater proportions of female participants (47/512, 9.18%) and participants aged 60 years or more (70/673, 10.40%) reported use of non-essential shops or services within 14 days post-vaccination. However, reported use of non-essential shops or services by male participants increased by more than female participants post-vaccination.

**Table 3.**
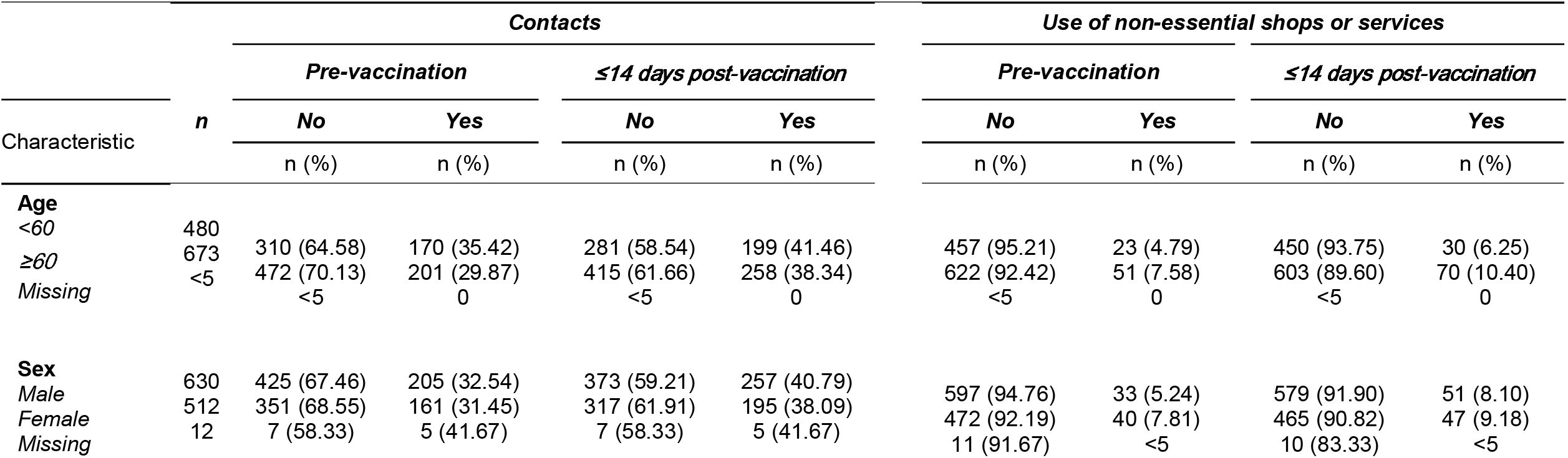
The frequency of reporting non-household contacts and use of non-essential shops or services over 7 days in pre-vaccination and post-vaccination surveys by participant characteristics (n = 1,154). The pre-vaccination survey and post-vaccination surveys asked participants to report activities from 2-9 February 2021 and 2-9 March 2021 respectively.

Table 4 presents the results of conditional logistic models for the within-individual effects of vaccination on non-household close contacts and use of non-essential shops or services. Odds of reporting non-household close contacts (OR 1.65, 95% CI 1.31-2.06, p=<0.001) and use of non-essential shops or services (OR 1.50, 95% CI 1.03-2.17, p=0.032) were higher within 14 days post-vaccination compared to pre-vaccination.

**Table 4.** Conditional logistic regression models for the effect of vaccinations status on contacts and use of non-essential shops or services.

| Vaccination status | n | Paired OR | Std. Err. | 95% CI | P-value |
| --- | --- | --- | --- | --- | --- |
| **Contacts** |  |  |  |  |  |
| ≤14 days post-vaccination | 352 | 1.65 | 0.19 | 1.31-2.06 | <0.001 |
| *Pre-vaccination* |  |  | - | - | - |
| **Use of non-essential shops or services** |  |  |  |  |  |
| ≤14 days post-vaccination | 125 | 1.50 | 0.28 | 1.03-2.17 | 0.032 |
| *Pre-vaccination* |  |  | - | - | - |

*Models adjusted for household structure with cluster-robust standard errors.*

Table 5 shows the results of stratified conditional logistic models examining heterogeneity of the within-individual effect of a change in vaccination status for contacts and use of non-essential shops or services by sociodemographic characteristics. All analyses compare time periods within 14 days post-vaccination to time periods pre-vaccination.

**Table 5.** Conditional logistic analyses stratified by sociodemographic characteristics for the effect of vaccination status on self-reported contacts and use of non-essential shops or services within 14 days of the first dose of a COVID-19 vaccine compared to pre-vaccination.

| Characteristic | Contacts |  |  |  |  | Use of non-essential shops or services |  |  |  |  |
| --- | --- | --- | --- | --- | --- | --- | --- | --- | --- | --- |
|  | n | Paired OR | Std. Err. | 95% CI | P-value | n | Paired OR | Std. Err. | 95% CI | P-value |
| **Sex** |  |  |  |  |  |  |  |  |  |  |
| Male | 198 | 1.71 | 0.25 | 1.28-2.29 | <0.001 | 60 | 1.86 | 0.51 | 1.08-3.19 | 0.025 |
| Female | 154 | 1.57 | 0.26 | 1.13-2.17 | 0.007 | 65 | 1.24 | 0.31 | 0.76-2.03 | 0.39 |
| **Age group** |  |  |  |  |  |  |  |  |  |  |
| <60 | 151 | 1.48 | 0.26 | 1.05-2.08 | 0.027 | 45 | 1.37 | 0.41 | 0.75-2.47 | 0.30 |
| ≥60 | 201 | 1.80 | 0.27 | 1.33-2.41 | <0.001 | 80 | 1.58 | 0.36 | 1.01-2.48 | 0.046 |

*Models adjusted for household structure with cluster-robust standard errors.*

After stratification by sex, greater odds of reporting non-household close contacts within the 14 days post-vaccination compared to pre-vaccination were consistent for both men (OR 1.71, 95% CI 1.28-2.29, p=<0.001) and women (OR 1.57, 95% CI 1.13-2.17, p=0.007). However, overlapping confidence intervals indicate a lack of effect heterogeneity by sex in the study population. There was evidence of effect of vaccination status on use of non-essential shops or services amongst male participants, who had greater odds of reporting these activities post-vaccination (OR 1.86 95% CI 1.14-3.32, p=0.015). Female participants were not at increased odds of reporting use of non-essential shops or services (OR 1.24, 95% CI 0.76-2.03, p=<0.39).

The odds of reporting non-household contacts and use of non-essential shops or services in those aged less than 60 years were respectively 1.48 (95% CI 1.05-2.08, p=0.027) and 1.37 (95% CI 0.75-2.47, p=0.30). Elevated odds were seen in those aged 60 years or more, with greater odds for both non-household close contacts (OR 1.80, 95% CI 1.33-2.41, p=<0.001) and use of non-essential shops or services (OR 1.58, 95% CI 1.01-2.48, p=0.046).

## DISCUSSION

Our findings indicate that within 14 days of their first dose of a COVID-19 vaccine, participants were more likely to report non-household close contacts (OR=1.65, 1.31-2.06) and visiting non-essential shops and services (OR=1.50, 1.03-2.17) compared to pre-vaccination. There was no substantial heterogeneity in the effect of vaccination status on non-household close contacts between men and women, but men had higher odds of reporting use of non-essential shops or services. Participants aged 60 years or more had greater odds of reporting both non-household close contacts and use of non-essential shops or services post-vaccination. Increased close contact with people outside of the household and in non-essential retail activities may contribute to the increased risk of infection seen within 14 days of a first dose of a COVID-19 vaccine (12,13), although directly measuring the risk of infection associated with these behaviours was outside the scope of this analysis.

The greatest effect was seen for contacts with people outside of the household. Few participants reported use of non-essential shops or services or social activities, leading to the exclusion of social activities from this analysis due to small group sizes. This is largely due to closure of public venues (e.g., non-essential retail or indoor dining venues) under national regulations in England and Wales at the time of the survey. Results from stratified analyses must also be interpreted with caution due to small subgroup sizes, which likely substantially reduced power for these analyses.

These results highlight the need to maintain protective behaviours within 14 days of a first dose of a COVID-19 vaccine while immunological protection builds. This could reduce the risk of infection in this time and is a relevant target for public health messaging. Although this analysis focused specifically on non-essential contacts and activities during the third national lockdown, it is possible that the effect of vaccination on activity levels at times when social distancing and other public health measures are relaxed may be even greater. Additionally, there is some evidence to suggest that older people are more likely to adopt or adhere to protective behaviours than younger people during pandemics (35). As most participants in this project were older than 50 years of age, our findings may be different or magnified in younger age groups. Our findings may also be of relevance for current and future booster doses of COVID-19 vaccines considering reported reductions in vaccine effectiveness over time (36-39). This may be of particular importance given recent evidence for immune evasion of both natural and vaccine-acquired immunity by the Omicron Variant of Concern (VoC) (40), the most dominant strain in the UK (41). Longer-term, quantitative data on behaviour related to vaccination could be used to parametrize and validate models of infectious disease dynamics used in public health decision making and policy (19,20), assisting public health planning in response to future outbreaks of infectious diseases.

While we found no differences in the effect of vaccination status between men and women, previous research has suggested that women may be more likely to adopt protective behaviours than men during pandemics, which is potentially related to increased perceived risk (35). A study examining adherence to public health guidance in the UK during the COVID-19 pandemic found that women reported making fewer trips outside of the home than men and less use of non-essential shops (42). Although our findings are supported by evidence that a reduction in protective behaviours may be seen after the introduction of vaccines during the COVID-19 pandemic (25, 43, 44), they are not consistent with those from a Virus Watch study using GPS location data to examine changes in travel distance pre- and post-the first dose of a COVID-19 vaccine during the third national lockdown (45). In this analysis, Nguyen et al. found no evidence for an increase in the rate of change in the distance travelled by participants post-vaccination in the 30 days before and after vaccination, suggesting that participants may not have altered behaviours leading to an increased distance travelled during this time. However, participants in the geolocation tracking arm of the study were aware that they were being monitored and so may have been more likely to modify their behaviour. Our results differ from those of Wright et al. (26), whose research showed that compliance with public health guidelines increased in both unvaccinated and vaccinated individuals from October 2020 to February 2021.

Context-specific psychological predictors of behaviour could account for this difference. A scoping review of 149 studies of behaviour change during pandemics (including the COVID-19 pandemic) found higher perceived risk predicted greater levels of adherence to a number of protective behaviours, including social distancing and avoidance of non-essential shopping (46). A number of contextual factors could have caused changes in perceived risk between late 2020 and early 2021. Perceived risk may have been greater during the national lockdown beginning in January 2021 at the peak of the second wave of COVID-19 cases (47). The rapid expansion of first the SARS-CoV-2 Alpha VoC (48) and then Delta VoC in early 2021 (49) may have also influenced perceived risk. Comparison of longer-term changes in compliance with guidelines or protective behaviours post-vaccination with changes over a shorter period of time may therefore not be appropriate.

Alternatively, perceived risk may contribute to the changes in behaviour seen in this analysis independently of vaccination status. Following the peak in January 2021, the number of people testing positive for COVID-19 decreased over February – March 2021 (50). Over time participants may have felt at less risk of COVID-19 and gradually increased their contacts and activities independently of a change in their vaccination status. As our analysis did not directly examine the perceived risk of infection further investigation of the relationship between psychological predictors of behaviour, vaccination and infectious disease dynamics is warranted.

A strength of the case-crossover design is that time-invariant confounders are controlled by self-matching. This is advantageous when studying complex outcomes such as behaviour as conditional logistic regression accounts for unmeasured confounders, which may be unknown or difficult to quantify. By selecting pre- and post-vaccination time periods to be within a period of time during which public health guidance and restrictions were in place, temporal and spatial variation in behaviours was likely minimised. The potential for information bias is minimised as survey questions remained the same over time, and self-matching means that the same individuals reported information for both pre- and post-vaccination time periods,

An important limitation of this analysis is the lack of representativeness of the study population with the general population in England and Wales. Participants who responded to surveys were predominantly of White ethnicity, living in the East or South East of England and living in less deprived environments with high household income. This is significant given the unequal burden of disease and mortality from COVID-19 in the UK population. People living in overcrowded households are at greater risk of SARS-CoV-2 infection (51), and rates of diagnosis and death are higher in people who live in more deprived areas and are from minority ethnic backgrounds (52). Furthermore, confounders (e.g., household income, IMD and region) may have varied over time but were only available at baseline. Self-reported contacts and activities may have been affected by recall bias, although this may have been minimised by survey timing (i.e. the following week). There was also the potential for social desirability bias, but this may have been reduced by survey responses being recorded online. Importantly, the infection risk of contacts and activities in this project could not be directly measured using survey responses – this limits inferences about the impact of changes in behaviour on infection risk and is recommended as a focus for future research.

This analysis provides quantitative evidence for an association between vaccination status and transmission-relevant behaviour using data gathered during the third national lockdown in England and Wales. Our findings suggest that changes in protective behaviours occur while immunological protection is building and may contribute to risk of infection in the 14 days after vaccination. It is possible that such an effect could exist with current and future booster doses of COVID-19 vaccines given reductions in vaccine effectiveness occurring over time. Interventions emphasising the need to maintain protective behaviours in the recently vaccinated could therefore reduce the risk of infection during these time periods.

## Data Availability

We aim to share aggregate data from this project on our website and via a "Findings so far" section on our website - https://ucl-virus-watch.net/. We also share some individual record level data on the Office of National Statistics Secure Research Service. In sharing the data we will work within the principles set out in the UKRI Guidance on best practice in the management of research data. Access to use of the data whilst research is being conducted will be managed by the Chief Investigators (ACH and RWA) in accordance with the principles set out in the UKRI guidance on best practice in the management of research data. We will put analysis code on publicly available repositories to enable their reuse.

## ABBREVIATIONS

COVID-19: *Coronavirus disease 2019*
SARS-CoV-2: *Severe acute respiratory syndrome coronavirus 2*
UK: *United Kingdom*
IMD: I*ndex of Multiple Deprivation*
VoC: *Variant of Concern*

## FIGURES

***Figure 1.*** Exclusion criteria for the analysis of the effect of vaccination status on contacts and activities.

